# Anatomy Segmentation in Laparoscopic Surgery: Comparison of Machine Learning and Human Expertise – An Experimental Study

**DOI:** 10.1101/2022.11.11.22282215

**Authors:** Fiona R. Kolbinger, Franziska M. Rinner, Alexander C. Jenke, Matthias Carstens, Stefanie Krell, Stefan Leger, Marius Distler, Jürgen Weitz, Stefanie Speidel, Sebastian Bodenstedt

## Abstract

**Background:** Lack of anatomy recognition represents a clinically relevant risk in abdominal surgery. Machine learning (ML) methods can help identify visible patterns and risk structures, however, their practical value remains largely unclear.

**Materials and Methods:** Based on a novel dataset of 13195 laparoscopic images with pixel-wise segmentations of eleven anatomical structures, we developed specialized segmentation models for each structure and combined models for all anatomical structures using two state-of-the-art model architectures (DeepLabv3 and SegFormer), and compared segmentation performance of algorithms to a cohort of 28 physicians, medical students, and medical laypersons using the example of pancreas segmentation.

**Results:** Mean Intersection-over-Union for semantic segmentation of intraabdominal structures ranged from 0.28 to 0.83 and from 0.23 to 0.77 for the DeepLabv3-based structure-specific and combined models, and from 0.31 to 0.85 and from 0.26 to 0.67 for the SegFormer-based structure-specific and combined models, respectively. Both the structure-specific and the combined DeepLabv3-based models are capable of near-real-time operation, while the SegFormer-based models are not. All four models outperformed at least 26 out of 28 human participants in pancreas segmentation.

**Conclusions:** These results demonstrate that ML methods have the potential to provide relevant assistance in anatomy recognition in minimally-invasive surgery in near-real-time. Future research should investigate the educational value and subsequent clinical impact of respective assistance systems.

**Highlights:**

- Machine learning models to reduce surgical risks that precisely identify 11 anatomical structures: abdominal wall, colon, intestinal vessels (inferior mesenteric artery and inferior mesenteric vein with their subsidiary vessels), liver, pancreas, small intestine, spleen, stomach, ureter and vesicular glands
- Large training dataset of 13195 real-world laparoscopic images with high-quality anatomy annotations
- Similar performance of individual segmentation models for each structure and combined segmentation models in identifying intraabdominal structures, and similar segmentation performance of DeepLabv3-based and SegFormer-based models
- DeepLabv3-based models are capable of near-real-time operation while SegFormer-based models are not, but SegFormer-based models outperform DeepLabv3-based models in terms of accuracy and generalizability
- All models outperformed at least 26 out of 28 human participants in pancreas segmentation, demonstrating their potential for real-time assistance in recognizing anatomical landmarks during minimally-invasive surgery.

## Introduction

Computer vision describes the computerized analysis of digital images aiming at the automation of human visual capabilities, most commonly using machine learning methods, in particular deep learning. This approach has transformed medicine in recent years, with successful applications including computer-aided diagnosis of colonic polyp dignity in endoscopy (1,2), detection of clinically actionable genetic alterations in histopathology (3), and melanoma detection in dermatology (4). Availability of large amounts of training data is the defining prerequisite for successful application of deep learning methods. With the establishment of laparoscopy as the gold standard for a variety of surgical procedures (5–8) and the increasing availability of computing resources, these concepts have gradually been applied to abdominal surgery. The overwhelming majority of research efforts in the field of Artificial Intelligence (AI)-based analysis of intraoperative surgical imaging data (i.e. video data from laparoscopic or open surgeries) has focused on classifying images with respect to the presence and/or location of previously annotated surgical instruments or anatomical structures (9–13) or on analysis of surgical proficiency (14–16) based on recorded procedures. However, almost all research endeavors in the field of computer vision in laparoscopic surgery have concentrated on preclinical stages and to date, no AI model based on intraoperative surgical imaging data could demonstrate a palpable clinical benefit (17,18). Among the studies closest to clinical application are recent works on identification of instruments and hepatobiliary anatomy during cholecystectomy for automated assessment of the critical view of safety (13), and on the automated segmentation of safe and unsafe preparation zones during cholecystectomy (19).

In surgery, patient outcome heavily depends on experience and performance of the surgical team (20,21). In a recent analysis of Human Performance Deficiencies in major cardiothoracic, vascular, abdominal transplant, surgical oncology, acute care, and general surgical operations, more than half of the cases with postoperative complications were associated with identifiable human error. Among these errors, lack of recognition (including misidentified anatomy) accounted for 18.8%, making it the most common Human Performance Deficiency overall (22). Examples of complications directly related to anatomical misperception are iatrogenic lesions to the ureter in gynecologic procedures (23) and pancreatic injuries during splenic flexure mobilization in colorectal surgery (24). While AI-based systems identifying anatomical risk and target structures would theoretically have the potential to alleviate this risk, limited availability and diversity of (annotated) laparoscopic image data drastically restrict the clinical potential of such applications in practice.

To advance and diversify the applications of computer vision in laparoscopic surgery, we have recently published the Dresden Surgical Anatomy Dataset (25), providing 13195 laparoscopic images with high-quality (26), expert-reviewed annotations of the presence and exact location of eleven intraabdominal anatomical structures: abdominal wall, colon, intestinal vessels (inferior mesenteric artery and inferior mesenteric vein with their subsidiary vessels), liver, pancreas, small intestine, spleen, stomach, ureter and vesicular glands. Here, we present the first study based on this dataset and present machine learning models to assist in precisely delineating anatomical structures, aiming to reduce surgical risks. Specifically, we evaluate automated detection and localization of organs and anatomical structures in laparoscopic view using two state-of-the-art model architectures: DeepLabv3 and SegFormer. To assess the clinical value of the presented machine learning models, we compare algorithm segmentation performance to that of humans using the example of delineation of the pancreas.

## Methods

### Patient cohort

Video data from 32 robot-assisted anterior rectal resections or rectal extirpations were gathered at the University Hospital Carl Gustav Carus Dresden between February 2019 and February 2021. All included patients had a clinical indication for the surgical procedure, recommended by an interdisciplinary tumor board. The procedures were performed using the da Vinci® Xi system (Intuitive Surgical, Sunnyvale, CA, USA) with a standard Da Vinci® Xi/X Endoscope with Camera (8 mm diameter, 30° angle, Intuitive Surgical, Sunnyvale, CA, USA, Item code 470057). Surgeries were recorded using the CAST system (Orpheus Medical GmBH, Frankfurt a.M., Germany). Each record was saved at a resolution of 1920 x 1080 pixels in MPEG-4 format.

All experiments were performed in accordance with the ethical standards of the Declaration of Helsinki and its later amendments. The local Institutional Review Board (ethics committee at the Technical University Dresden) reviewed and approved this study (approval number: BO-EK-140032021). The trial was registered on clinicaltrials.gov (trial registration ID: NCT05268432). Written informed consent to laparoscopic image data acquisition, data annotation, data analysis, and anonymized data publication was obtained from all participants. Before publication, all data was anonymized according to the general data protection regulation of the European Union.

### Patient cohort

Video data from 32 robot-assisted anterior rectal resections or rectal exstirpations were gathered at the University Hospital Carl Gustav Carus Dresden between February 2019 and February 2021. All included patients had a clinical indication for the surgical procedure, recommended by an interdisciplinary tumor board. Patients were not specifically selected with respect to demographic or physical parameters (i.e. age, sex, body-mass index, comorbidities, previous surgical procedures) or disease-specific criteria (i.e. indication, disease stage). Respective details of the underlying patient cohort have been published previously (25). The procedures were performed using the da Vinci® Xi system (Intuitive Surgical, Sunnyvale, CA, USA) with a standard Da Vinci® Xi/X Endoscope with Camera (8 mm diameter, 30° angle, Intuitive Surgical, Sunnyvale, CA, USA, Item code 470057). Surgeries were recorded using the CAST system (Orpheus Medical GmBH, Frankfurt a.M., Germany). Each record was saved at a resolution of 1920 x 1080 pixels in MPEG-4 format.

All experiments were performed in accordance with the ethical standards of the Declaration of Helsinki and its later amendments. The local Institutional Review Board (ethics committee at the Technical University Dresden) reviewed and approved this study (approval number: BO-EK-140032021). The trial was registered on clinicaltrials.gov (trial registration ID: NCT05268432). Written informed consent to laparoscopic image data acquisition, data annotation, data analysis, and anonymized data publication was obtained from all participants. Before publication, all data was anonymized according to the general data protection regulation of the European Union.

### Dataset

Based on the full-length surgery recordings and respective temporal annotations of organ visibility, individual image frames were extracted and annotated as described previously (25). In brief, three independent annotators with substantial experience in robot-assisted rectal surgery created pixel-wise annotations, which were subsequently reviewed by a surgeon with 4 years of experience in robot-assisted rectal surgery. A detailed description of the annotation process including underlying annotation protocols as well as analyses of annotator agreement and technical parameters has been published previously (25). To guarantee real-world applicability of machine learning models trained on the dataset, images with perturbations such as blurring due to camera movements, soiling of the lens, and presence of blood or smoke were not specifically excluded. However, the annotation protocols advised annotators to only annotate structures in soiled and blurry images if the respective structures were clearly delineable. The resulting Dresden Surgical Anatomy Dataset comprises 13195 distinct images with pixel-wise segmentations of eleven anatomical structures: abdominal wall, colon, intestinal vessels (inferior mesenteric artery and inferior mesenteric vein with their subsidiary vessels), liver, pancreas, small intestine, spleen, stomach, ureter and vesicular glands. Moreover, the dataset comprises binary annotations of the presence of each of these organs for each image. The dataset is publicly available via the following link: https://doi.org/10.6084/m9.figshare.21702600.

For machine learning purposes, the Dresden Surgical Anatomy Dataset was split into training, validation, and test data as follows (Figure 1):

**Figure 1:**
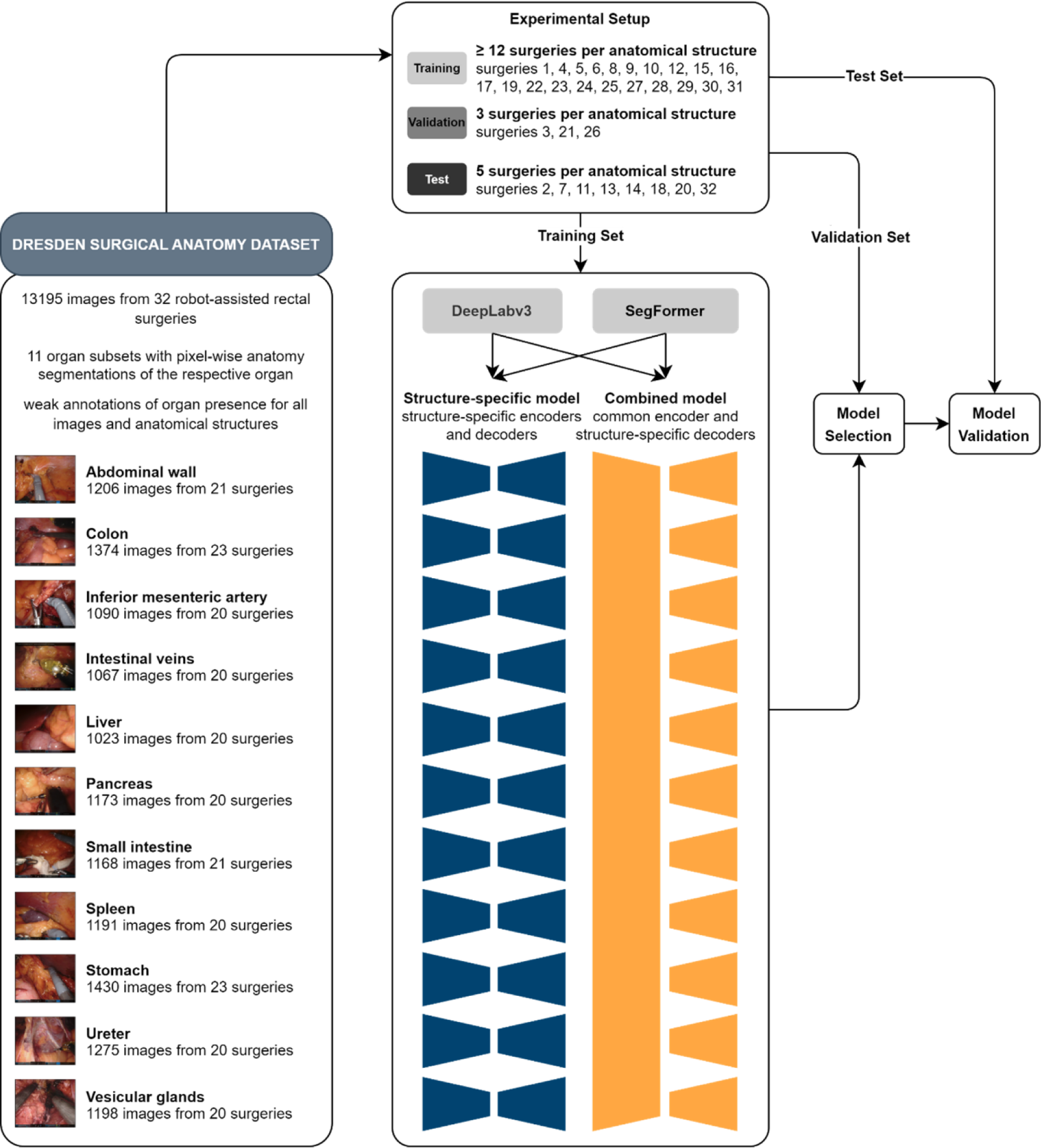
Schematic illustration of the structure-specific and combined machine learning models used for semantic segmentation. The Dresden Surgical Anatomy Dataset was split into a training, a validation, and a test set. For spatial segmentation, two sets of machine learning models – a structure-specific model with individual encoders and decoders, and a combined model with a common encoder and structure-specific decoders – were trained for DeepLabv3-based and SegFormer-based model architectures.

— Training set (at least 12 surgeries per anatomical structure): surgeries 1, 4, 5, 6, 8, 9, 10, 12, 15, 16, 17, 19, 22, 23, 24, 25, 27, 28, 29, 30, 31.
— Validation set (3 surgeries per anatomical structure): surgeries 3, 21, 26.
— Test set (5 surgeries per anatomical structure): surgeries 2, 7, 11, 13, 14, 18, 20, 32.

This split is proposed for future works using the Dresden Surgical Anatomy Dataset to reproduce the variance of the entire dataset within each subset, and to ensure comparability regarding clinical variables between the training, the validation, and the test set. Surgeries for the test set were selected to minimize variance regarding the number of frames over the segmented classes. Out of the remaining surgeries, the validation set was separated from the training set using the same criterion.

### Structure-specific semantic segmentation models

To segment each anatomical structure, a separate convolutional neural network for segmentation of individual structures was trained. Specifically, we trained and compared two different architectures: a Deeplabv3 (27) model with a ResNet50 backbone with default PyTorch pretraining on the COCO dataset (28), and a SegFormer (29) model pretrained on the Cityscapes dataset (30). The networks were trained using cross-entropy loss and the AdamW optimizer (31) for 100 epochs with a starting learning rate of 10^-4^ and a linear learning rate scheduler decreasing the learning rate by 0.9 every 10 epochs. For data augmentation, we applied random scaling and rotation, as well as brightness and contrast adjustments. The final model for each organ was selected via the Intersection-over-Union (IoU, Supplementary Figure 1) on the validation dataset and evaluated using the Dresden Surgical Anatomy Dataset with the abovementioned training-validation-test split (Figure 1).

Segmentation performance was assessed using F1 score, IoU, precision, recall, and specificity on the test folds. These parameters are commonly used technical measures of prediction exactness, ranging from 0 (least exact prediction) to 1 (entirely correct prediction without any misprediction, Supplementary Figure 1).

### Combined semantic segmentation models

A convolutional neural network with a common encoder and eleven decoders for combined segmentation of the eleven anatomical structures was trained. As for the structure-specific models, DeepLabv3-based (27) and SegFormer-based (29) models were used. For DeepLabv3, a shared ResNet50 backbone with default PyTorch pretraining on the COCO dataset (28) was used. For each class, a DeepLabv3 decoder was then run on the features extracted from a given image by the backbone. Similarly, for SegFormer, an encoder, pretrained on the Cityscapes dataset (30), was combined with eleven decoders.

As the images are only annotated for binary classes, the loss is only calculated for every pixel in images, in which the structure associated with the current decoder is annotated. For images, in which the associated class is not annotated, only the pixels that are annotated as belonging to another class are included in the loss, e.g., pixels that were annotated as belonging to the class “liver” can be used as negative examples for the class “pancreas”. The remaining training procedure was identical to the structure-specific model. The models were trained and evaluated using the Dresden Surgical Anatomy Dataset with the abovementioned training-validation-test split (Figure 1).

Segmentation performance was assessed using F1 score, IoU, precision, recall, and specificity on the test folds.

### Evaluation of the semantic segmentation models on an external dataset

To explore generalizability, structure-specific and combined models based on both architectures (DeepLabv3 and SegFormer) were deployed to laparoscopic image data from the publicly available LapGyn4 dataset (32). Models were separately deployed for full-scene segmentations and their performance was visually compared.

### Comparative evaluation of algorithmic and human performance

To determine the clinical potential of automated segmentation of anatomical risk structures, the segmentation performance of 28 humans was compared to that of the structure-specific and the combined semantic segmentation models using the example of the pancreas. The local Institutional Review Board (ethics committee at the Technical University Dresden) reviewed and approved this study (approval number: BO-EK-566122021). All participants provided written informed consent to anonymous study participation, data acquisition and analysis, and publication. In total, 28 participants (physician and non-physician medical staff, medical students, and medical laypersons) marked the pancreas in 35 images from the Dresden Surgical Anatomy Dataset (25) with bounding boxes. These images originated from 26 different surgeries, and the pancreas was visible in 16 of the 35 images. Each of the previously selected 35 images was shown once, the order being arbitrarily chosen but identical for all participants. The open-source annotation software Computer Vision Annotation Tool (CVAT) was used for annotations. In cases where the pancreas was seen in multiple, non-connected locations in the image, participants were asked to create separate bounding boxes for each area.

Based on the structure-specific and the combined semantic segmentation models, axis-aligned bounding boxes marking the pancreas were generated in the 35 images from the pixel-wise segmentation. To guarantee that the respective images were not part of the training data, four-fold cross validation was used, i.e., the origin surgeries were split into four equal-sized batches, and algorithms were trained on three batches that did not contain the respective origin image before being applied to segmentation.

To compare human and algorithm performance, the bounding boxes created by each participant and the structure-specific as well as the combined semantic segmentation models were compared to bounding boxes derived from the Dresden Surgical Anatomy Dataset, which were defined as ground truth. IoU between the manual or automatic bounding box and the ground truth was used to compare segmentation accuracy.

## Data and Code Availability

### Data Availability

The Dresden Surgical Anatomy Dataset is publicly available via the following link: https://doi.org/10.6084/m9.figshare.21702600. All other data generated and analyzed during the current study are available from the corresponding authors on reasonable request. To gain access, data requestors will need to sign a data access agreement.

### Code Availability

The most relevant scripts used for dataset compilation are publicly available via the following link: https://zenodo.org/record/6958337#.YzsBdnZBzOg. The code used for segmentation algorithms is available at https://gitlab.com/nct_tso_public/anatomy-recognition-dsad.

## Results

***Machine Learning-based anatomical structure segmentation in structure-specific models*** Structure-specific multi-layer convolutional neural networks (Figure 1) based on two different semantic segmentation architectures termed DeepLabv3 and SegFormer, were trained to segment the abdominal wall, the colon, intestinal vessels (inferior mesenteric artery and inferior mesenteric vein with their subsidiary vessels), the liver, the pancreas, the small intestine, the spleen, the stomach, the ureter, and vesicular glands (Supplementary Table 1). Table 1 displays technical metrics of overlap between the annotated ground truth and the model predictions (mean F1 score, IoU, precision, recall, and specificity) for individual anatomical structures as predicted by the structure-specific algorithms on the test data.

**Table 1:** Summary of performance metrics for anatomical structure segmentation using DeepLabv3-based (a) and SegFormer-based (b) structure-specific models on the test dataset. For each metric, mean and standard deviation are displayed.

| a | **Anatomical structure** | **F1 score** | **IoU** | **Precision** | **Recall** | **Specificity** |
| --- | --- | --- | --- | --- | --- | --- |
|  | Abdominal wall | 0.90 ± 0.10 | 0.83 ± 0.14 | 0.89 ± 0.14 | 0.93 ± 0.07 | 0.97 ± 0.04 |
|  | Colon | 0.79 ± 0.20 | 0.69 ± 0.22 | 0.80 ± 0.21 | 0.82 ± 0.21 | 0.97 ± 0.05 |
|  | Inferior mesenteric artery | 0.54 ± 0.26 | 0.41 ± 0.22 | 0.55 ± 0.25 | 0.67 ± 0.33 | 0.99 ± 0.01 |
|  | Intestinal veins | 0.54 ± 0.33 | 0.44 ± 0.29 | 0.70 ± 0.26 | 0.56 ± 0.36 | 1.00 ± 0.00 |
|  | Liver | 0.80 ± 0.23 | 0.71 ± 0.25 | 0.85 ± 0.21 | 0.81 ± 0.24 | 0.98 ± 0.03 |
|  | Pancreas | 0.37 ± 0.32 | 0.28 ± 0.27 | 0.59 ± 0.37 | 0.37 ± 0.36 | 1.00 ± 0.01 |
|  | Small intestine | 0.87 ± 0.14 | 0.80 ± 0.18 | 0.87 ± 0.16 | 0.91 ± 0.15 | 0.97 ± 0.04 |
|  | Spleen | 0.79 ± 0.23 | 0.69 ± 0.24 | 0.74 ± 0.22 | 0.90 ± 0.24 | 0.99 ± 0.01 |
|  | Stomach | 0.71 ± 0.24 | 0.60 ± 0.25 | 0.65 ± 0.25 | 0.89 ± 0.21 | 0.98 ± 0.02 |
|  | Ureter | 0.47 ± 0.30 | 0.36 ± 0.25 | 0.53 ± 0.28 | 0.57 ± 0.39 | 1.00 ± 0.00 |
|  | Vesicular glands | 0.40 ± 0.25 | 0.28 ± 0.21 | 0.37 ± 0.28 | 0.62 ± 0.35 | 0.97 ± 0.03 |
| b | **Anatomical structure** | **F1 score** | **IoU** | **Precision** | **Recall** | **Specificity** |
|  | Abdominal wall | 0.91 ± 0.11 | 0.85 ± 0.15 | 0.90 ± 0.14 | 0.94 ± 0.09 | 0.98 ± 0.03 |
|  | Colon | 0.77 ± 0.21 | 0.66 ± 0.22 | 0.73 ± 0.22 | 0.87 ± 0.22 | 0.95 ± 0.07 |
|  | Inferior mesenteric artery | 0.60 ± 0.23 | 0.46 ± 0.21 | 0.58 ± 0.25 | 0.73 ± 0.29 | 0.99 ± 0.01 |
|  | Intestinal veins | 0.65 ± 0.25 | 0.52 ± 0.24 | 0.62 ± 0.27 | 0.76 ± 0.27 | 1.00 ± 0.00 |
|  | Liver | 0.83 ± 0.21 | 0.75 ± 0.24 | 0.82 ± 0.23 | 0.88 ± 0.18 | 0.98 ± 0.03 |
|  | Pancreas | 0.47 ± 0.32 | 0.37 ± 0.28 | 0.61 ± 0.36 | 0.48 ± 0.36 | 0.99 ± 0.01 |
|  | Small intestine | 0.89 ± 0.13 | 0.83 ± 0.17 | 0.87 ± 0.16 | 0.95 ± 0.10 | 0.97 ± 0.04 |
|  | Spleen | 0.85 ± 0.19 | 0.78 ± 0.21 | 0.80 ± 0.19 | 0.95 ± 0.16 | 1.00 ± 0.01 |
|  | Stomach | 0.75 ± 0.27 | 0.66 ± 0.28 | 0.76 ± 0.25 | 0.82 ± 0.29 | 0.99 ± 0.01 |
|  | Ureter | 0.58 ± 0.27 | 0.46 ± 0.24 | 0.53 ± 0.26 | 0.74 ± 0.32 | 0.99 ± 0.01 |
|  | Vesicular glands | 0.43 ± 0.26 | 0.31 ± 0.22 | 0.40 ± 0.28 | 0.63 ± 0.35 | 0.97 ± 0.03 |

Out of the analyzed segmentation models based on DeepLabv3, performance was lowest for vesicular glands (mean IoU: 0.28 ± 0.21), the pancreas (mean IoU: 0.28 ± 0.27), and the ureter (mean IoU: 0.36 ± 0.25), while excellent predictions were achieved for the abdominal wall (mean IoU: 0.83 ± 0.14) and the small intestine (mean IoU: 0.80 ± 0.18) (Supplementary Figure 1). In segmentation of the pancreas, the ureter, vesicular glands and intestinal vessel structures, there was a relevant proportion of images with no detection or no overlap between prediction and ground truth, while for all remaining anatomical structures, this proportion was minimal (Figure 2a). While the images, in which the highest IoUs were observed, mostly displayed large organ segments that were clearly visible (Figure 2 b), the images with the lowest IoU were of variable quality with confounding factors such as blood, smoke, soiling of the endoscope lens, or pictures blurred by camera shake (Figure 2 c). While overall segmentation performance of both architectures was similar for structure-specific models, SegFormer-based models showed a trend towards better performance than DeepLabv3-based models in segmentation of the pancreas, the spleen, and the ureter (Table 1, Figure 2, Supplementary Figure 2).

**Figure 2:**
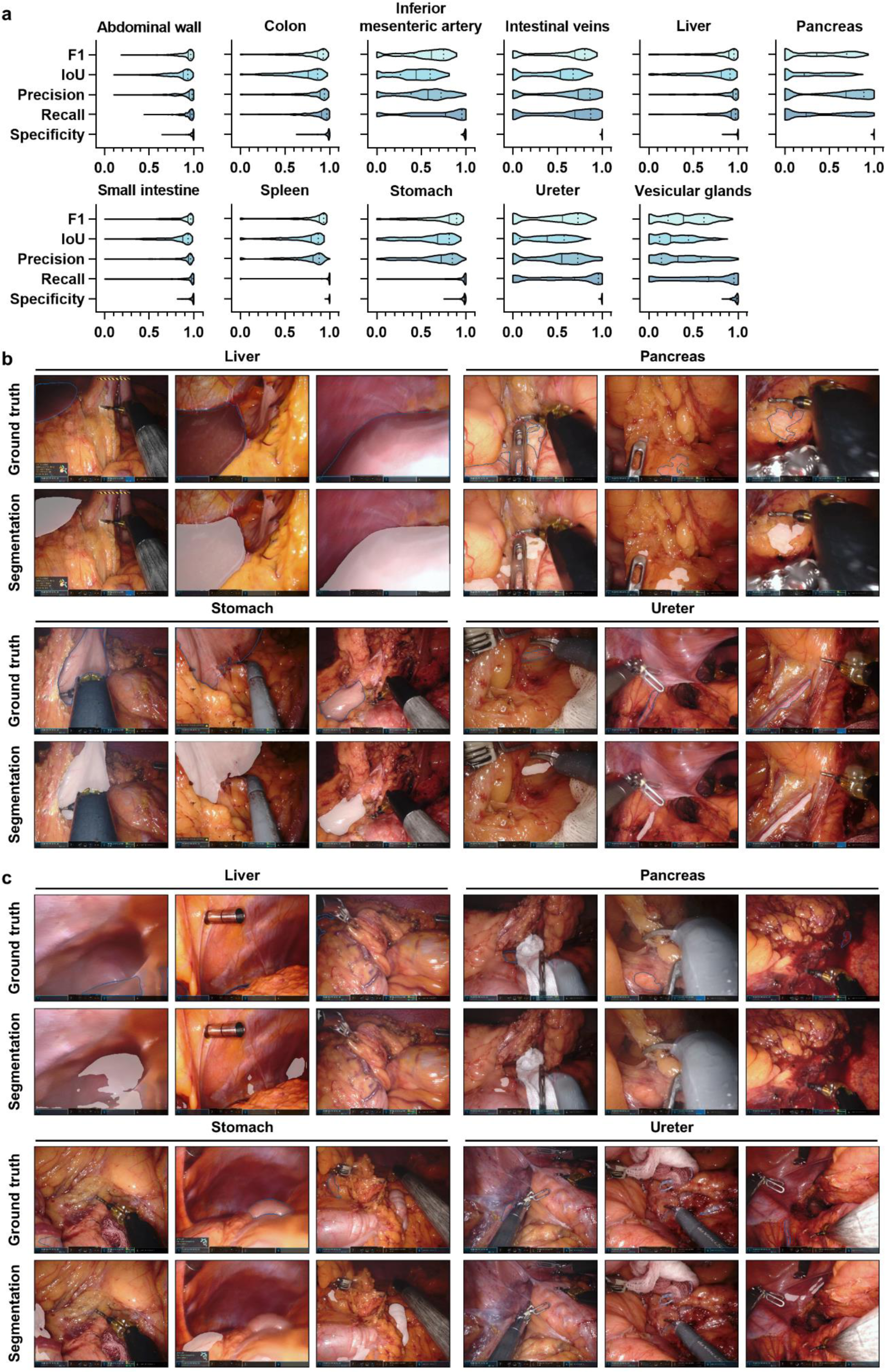
Pixel-wise organ segmentation with DeepLabv3-based structure-specific models trained on the respective organ subsets of the Dresden Surgical Anatomy Dataset. **(a)** Violin plot illustrations of performance metrics for DeepLabv3-based structure-specific segmentation models on the test dataset. The median and quartiles are illustrated as solid and dashed lines, respectively. **(b)** Example images from the test dataset with the highest IoUs for liver, pancreas, stomach, and ureter segmentation with DeepLabv3-based structure-specific segmentation models. Ground truth is displayed as blue line (upper panel), model segmentations are displayed as white overlay (lower panel). **(c)** Example images from the test dataset with the lowest IoUs for liver, pancreas, stomach, and ureter segmentation with DeepLabv3-based structure-specific segmentation models. Ground truth is displayed as blue line (upper panel), model segmentations are displayed as white overlay (lower panel).

To determine the models’ capabilities to operate in real-time (frame rates of > 20 frames per second), we determined their inference times per image. For the DeepLabv3-based structure-specific models, inference on a single image with a resolution of 640 x 512 pixels required, on average, 28 ms on an Nvidia A5000, resulting in a frame rate of almost 36 frames per second. In contrast, the SegFormer-based structure-specific semantic segmentation models operated considerably slower at an inference time of 53 ms per image, resulting in a frame rate of 18 frames per second. This runtime includes one decoder, meaning that only the segmentation for one anatomical class (organ or structure) is included.

### Machine Learning-based anatomical structure segmentation in combined models

In contrast to structure-specific models, models with a mutual encoder and organ-specific decoders could facilitate the identification of multiple organs at once, with the potential benefit of faster operation for multiple classes instead of sequential operation of several class-specific models. Therefore, combined models for both semantic segmentation architectures – DeepLabv3 and Segformer – were trained using annotated images from the Dresden Surgical Anatomy Dataset across anatomical structure classes (Figure 1, Supplementary Table 2). Table 2 displays mean F1 score, IoU, precision, recall, and specificity for anatomical structure segmentation in the combined model.

**Table 2:**
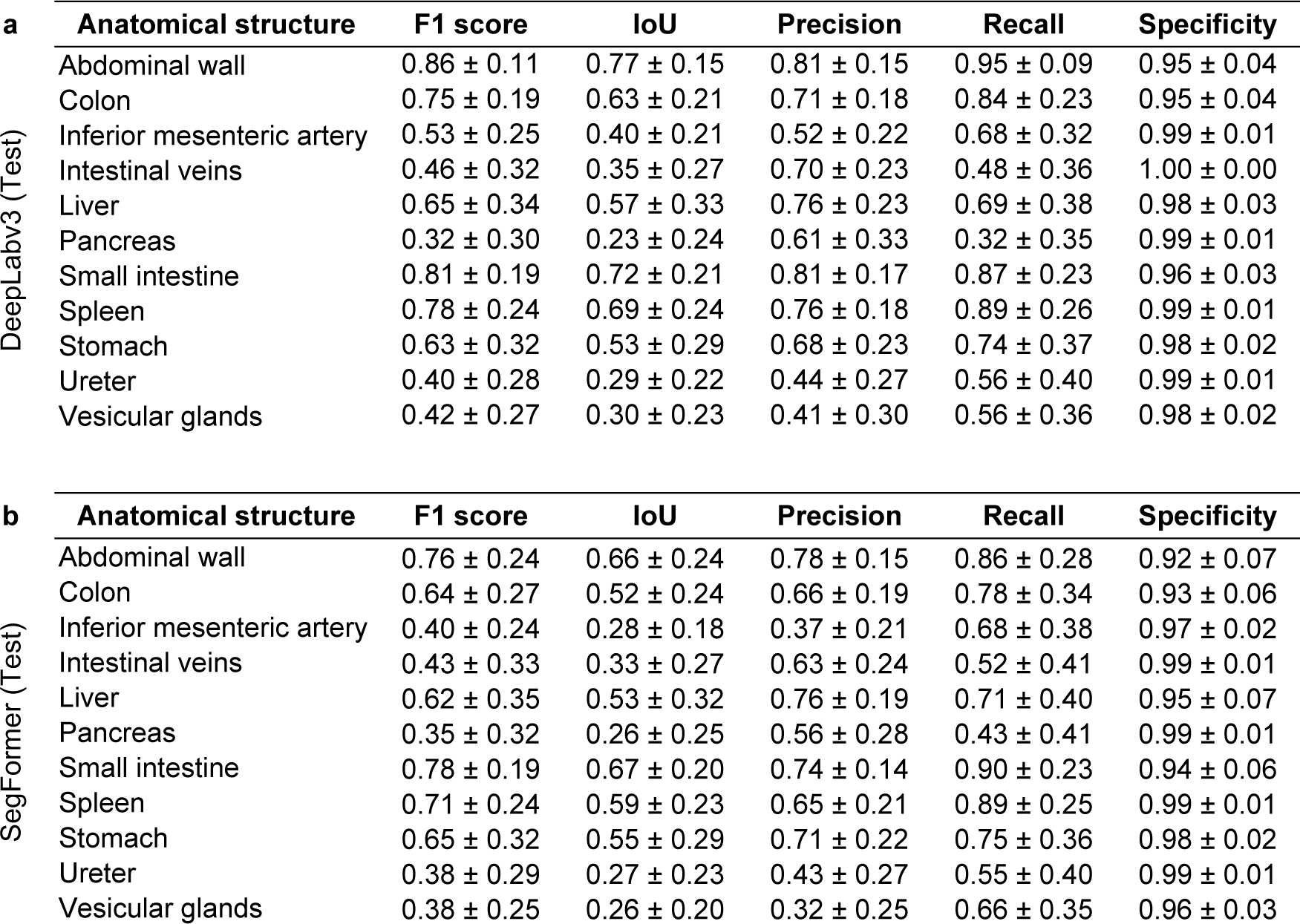
Summary of performance metrics for anatomical structure segmentation using the DeepLabv3-based (a) and SegFormer-based (b) combined models (common encoder with structure-specific decoders) on the test dataset. For each metric, mean and standard deviation are displayed.

| a | Anatomical structure | F1 score | IoU | Precision | Recall | Specificity |
| --- | --- | --- | --- | --- | --- | --- |
|  | Abdominal wall | 0.86 ± 0.11 | 0.77 ± 0.15 | 0.81 ± 0.15 | 0.95 ± 0.09 | 0.95 ± 0.04 |
|  | Colon | 0.75 ± 0.19 | 0.63 ± 0.21 | 0.71 ± 0.18 | 0.84 ± 0.23 | 0.95 ± 0.04 |
|  | Inferior mesenteric artery | 0.53 ± 0.25 | 0.40 ± 0.21 | 0.52 ± 0.22 | 0.68 ± 0.32 | 0.99 ± 0.01 |
|  | Intestinal veins | 0.46 ± 0.32 | 0.35 ± 0.27 | 0.70 ± 0.23 | 0.48 ± 0.36 | 1.00 ± 0.00 |
|  | Liver | 0.65 ± 0.34 | 0.57 ± 0.33 | 0.76 ± 0.23 | 0.69 ± 0.38 | 0.98 ± 0.03 |
|  | Pancreas | 0.32 ± 0.30 | 0.23 ± 0.24 | 0.61 ± 0.33 | 0.32 ± 0.35 | 0.99 ± 0.01 |
|  | Small intestine | 0.81 ± 0.19 | 0.72 ± 0.21 | 0.81 ± 0.17 | 0.87 ± 0.23 | 0.96 ± 0.03 |
|  | Spleen | 0.78 ± 0.24 | 0.69 ± 0.24 | 0.76 ± 0.18 | 0.89 ± 0.26 | 0.99 ± 0.01 |
|  | Stomach | 0.63 ± 0.32 | 0.53 ± 0.29 | 0.68 ± 0.23 | 0.74 ± 0.37 | 0.98 ± 0.02 |
|  | Ureter | 0.40 ± 0.28 | 0.29 ± 0.22 | 0.44 ± 0.27 | 0.56 ± 0.40 | 0.99 ± 0.01 |
|  | Vesicular glands | 0.42 ± 0.27 | 0.30 ± 0.23 | 0.41 ± 0.30 | 0.56 ± 0.36 | 0.98 ± 0.02 |
| b | Anatomical structure | F1 score | IoU | Precision | Recall | Specificity |
|  | Abdominal wall | 0.76 ± 0.24 | 0.66 ± 0.24 | 0.78 ± 0.15 | 0.86 ± 0.28 | 0.92 ± 0.07 |
|  | Colon | 0.64 ± 0.27 | 0.52 ± 0.24 | 0.66 ± 0.19 | 0.78 ± 0.34 | 0.93 ± 0.06 |
|  | Inferior mesenteric artery | 0.40 ± 0.24 | 0.28 ± 0.18 | 0.37 ± 0.21 | 0.68 ± 0.38 | 0.97 ± 0.02 |
|  | Intestinal veins | 0.43 ± 0.33 | 0.33 ± 0.27 | 0.63 ± 0.24 | 0.52 ± 0.41 | 0.99 ± 0.01 |
|  | Liver | 0.62 ± 0.35 | 0.53 ± 0.32 | 0.76 ± 0.19 | 0.71 ± 0.40 | 0.95 ± 0.07 |
|  | Pancreas | 0.35 ± 0.32 | 0.26 ± 0.25 | 0.56 ± 0.28 | 0.43 ± 0.41 | 0.99 ± 0.01 |
|  | Small intestine | 0.78 ± 0.19 | 0.67 ± 0.20 | 0.74 ± 0.14 | 0.90 ± 0.23 | 0.94 ± 0.06 |
|  | Spleen | 0.71 ± 0.24 | 0.59 ± 0.23 | 0.65 ± 0.21 | 0.89 ± 0.25 | 0.99 ± 0.01 |
|  | Stomach | 0.65 ± 0.32 | 0.55 ± 0.29 | 0.71 ± 0.22 | 0.75 ± 0.36 | 0.98 ± 0.02 |
|  | Ureter | 0.38 ± 0.29 | 0.27 ± 0.23 | 0.43 ± 0.27 | 0.55 ± 0.40 | 0.99 ± 0.01 |
|  | Vesicular glands | 0.38 ± 0.25 | 0.26 ± 0.20 | 0.32 ± 0.25 | 0.66 ± 0.35 | 0.96 ± 0.03 |

The performance of the combined model based on DeepLabv3 was overall similar to that of structure-specific models (Table 1), with highest segmentation performance for the abdominal wall (mean IoU: 0.77 ± 0.15) and the small intestine (mean IoU: 0.72 ± 0.21), and the lowest performance for the pancreas (mean IoU: 0.23 ± 0.29), the ureter (IoU: 0.29 ± 0.22) and vesicular glands (IoU: 0.30 ± 0.23) (Supplementary Figure 1). In comparison to the respective structure-specific models, the combined DeepLabv3-based model performed notably weaker in liver segmentation, while performance for the other anatomical structures was similar. The proportion of images for which the combined DeepLabv3-based model could not create a prediction or for which predictions showed no overlap with the ground truth at all was largest in the ureter, the pancreas, the stomach, the abdominal vessel structures, and the vesicular glands (Figure 3a). Similar to the DeepLabv3-based structure-specific models, trends towards an impact of segment size, uncommon angles of vision, endoscope lens soiling, blurry images, and presence of blood or smoke were seen when comparing image quality of well-predicted images (Figure 3 b) to images with poor or no prediction (Figure 3 c). Similar to the structure-specific models, segmentation performance of the SegFormer-based combined segmentation model was, overall, similar to that of DeepLabv3-based models. For segmentation of the spleen, there was a trend towards weaker performance of SegFormer-based models than DeepLabv3-based combined models (Table 2, Figure 3, Supplementary Figure 3).

**Figure 3:**
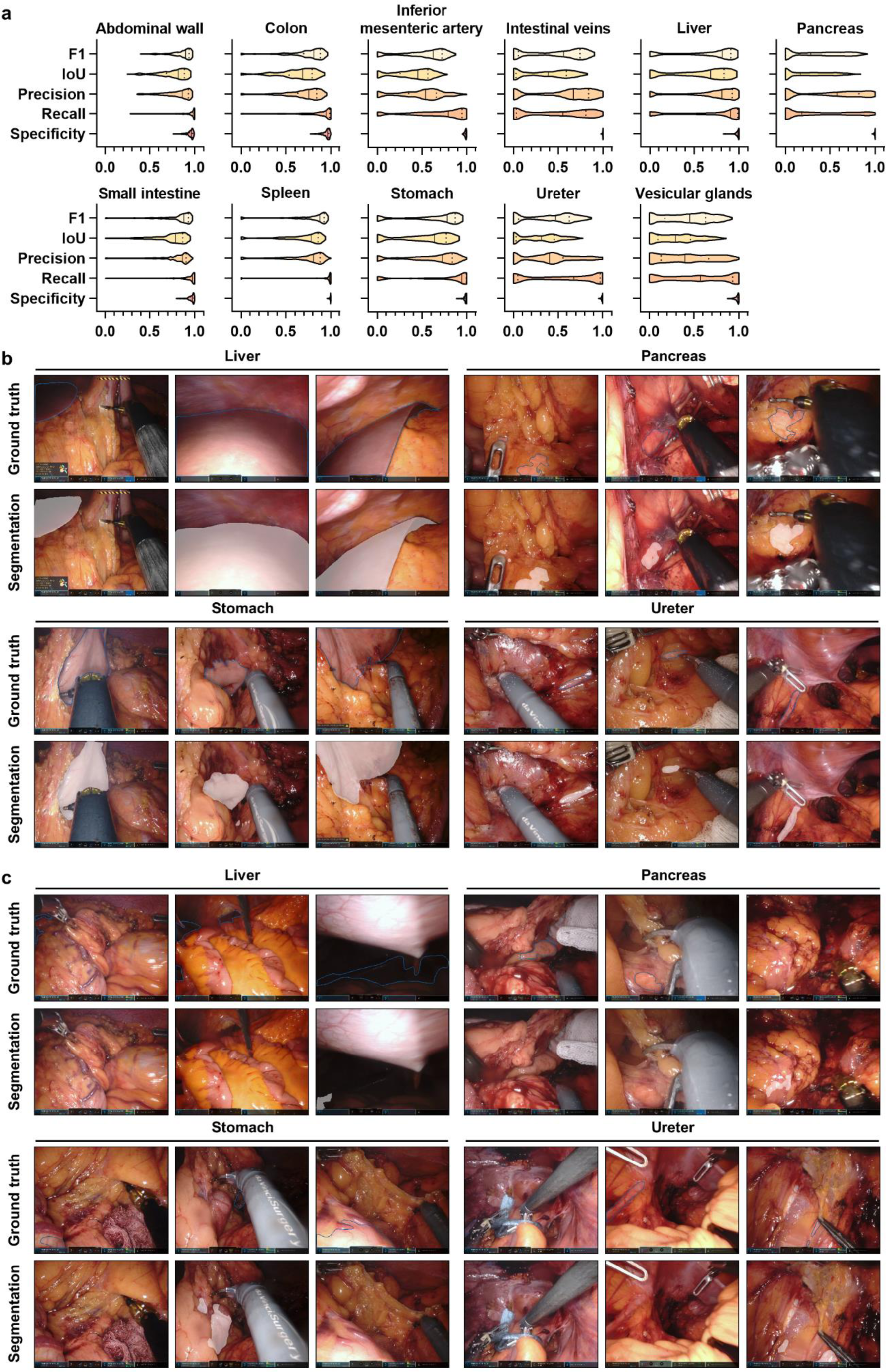
Pixel-wise organ segmentation with the DeepLabv3-based combined model trained on the Dresden Surgical Anatomy Dataset across anatomical structure classes with a common encoder and structure-specific decoders. **(a)** Violin plot illustrations of performance metrics for the DeepLabv3-based combined segmentation model on the test dataset. The median and quartiles are illustrated as solid and dashed lines, respectively. **(b)** Example images from the test dataset with the highest IoUs for liver, pancreas, stomach, and ureter segmentation with the DeepLabv3-based combined segmentation model. Ground truth is displayed as blue line (upper panel), model segmentations are displayed as white overlay (lower panel). **(c)** Example images from the test dataset with the lowest IoUs for liver, pancreas, stomach, and ureter segmentation with the DeepLabv3-based combined segmentation model. Ground truth is displayed as blue line (upper panel), model segmentations are displayed as white overlay (lower panel).

For the DeepLabv3-based combined models, inference on a single image with a resolution of 640 x 512 pixels required, on average, 71 ms on an Nvidia A5000, resulting in a frame rate of about 14 frames per second. As for structure-specific models of both architectures, SegFormer-based combined semantic segmentation models operated considerably slower at an inference time of 102 ms per image, resulting in a frame rate of about 10 frames per second. This runtime includes all 11 decoders, meaning that segmentations for all anatomical classes (organs or structures) are included.

### Performance of machine learning models on an external laparoscopic image dataset

To evaluate model robustness on an external dataset, we deployed the different organ segmentation models onto the publicly available LapGyn4 dataset (32) and qualitatively compared their performance. Overall, the combined models better reflected true anatomical constellations than the structure-specific models that generally lacked specificity. With respect to model architecture, the SegFormer-based segmentations were considerably more robust than the DeepLabv3-based models. Common mispredictions included confusion of liver and spleen, misinterpretation of organs that were not part of the training dataset (i.e. the gallbladder), and poor segmentation performance on less common images (i.e. extreme close-ups) (Figure 4).

**Figure 4:**
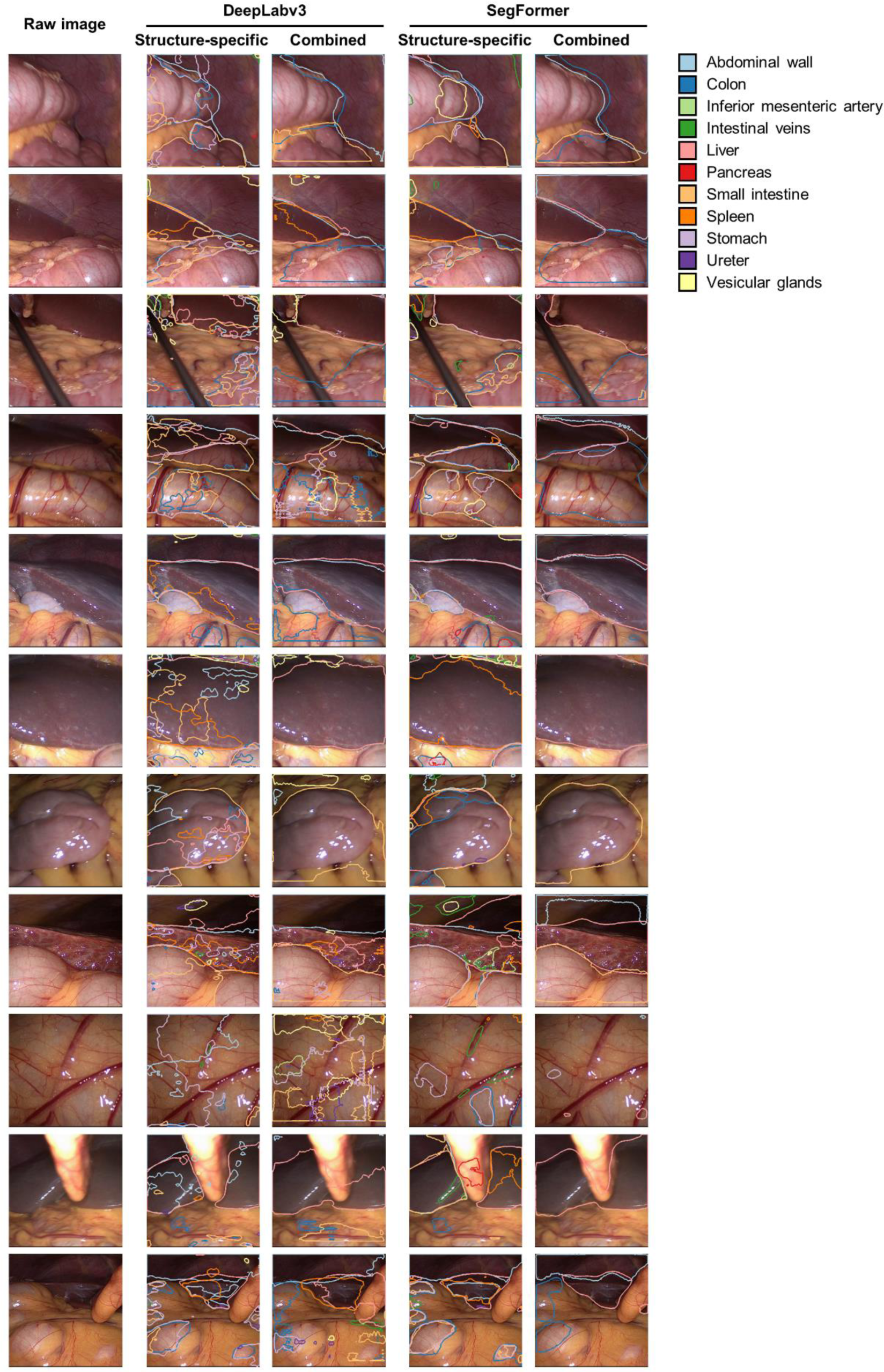
Comparison of DeepLabv3-based and Segformer-based structure-specific and combined segmentation model performance on an external laparoscopic image dataset (LapGyn4). Models were deployed to the publicly available LapGyn4 dataset of non-semantically segmented images from gynecological procedures in conventional laparoscopic technique. Model segmentations for each organ are displayed. For the structure-specific models, segmentations of the eleven individual segmentation models are overlayed in one image. Figure shows representative images from the dataset.

In summary, the SegFormer-based combined semantic segmentation model resulted in robust segmentations reproducing the true underlying anatomy. The remaining segmentation models provided substantially less specific and less robust segmentation outputs on the external dataset.

### Performance of machine learning models in relation to human performance

To approximate the clinical value of the previously described algorithms for anatomical structure segmentation, the performances of the DeepLabv3-based and SegFormer-based structure-specific and the combined models were compared to that of a cohort of 28 physicians, medical students, and persons with no medical background (Figure 5a), and different degrees of experience in laparoscopic surgery (Figure 5 b). A vulnerable anatomical structure (24) with – measured by classical metrics of overlap (Tables 1 and 2) – comparably weak segmentation performance of the trained algorithms, the pancreas was selected as an example.

**Figure 5:**
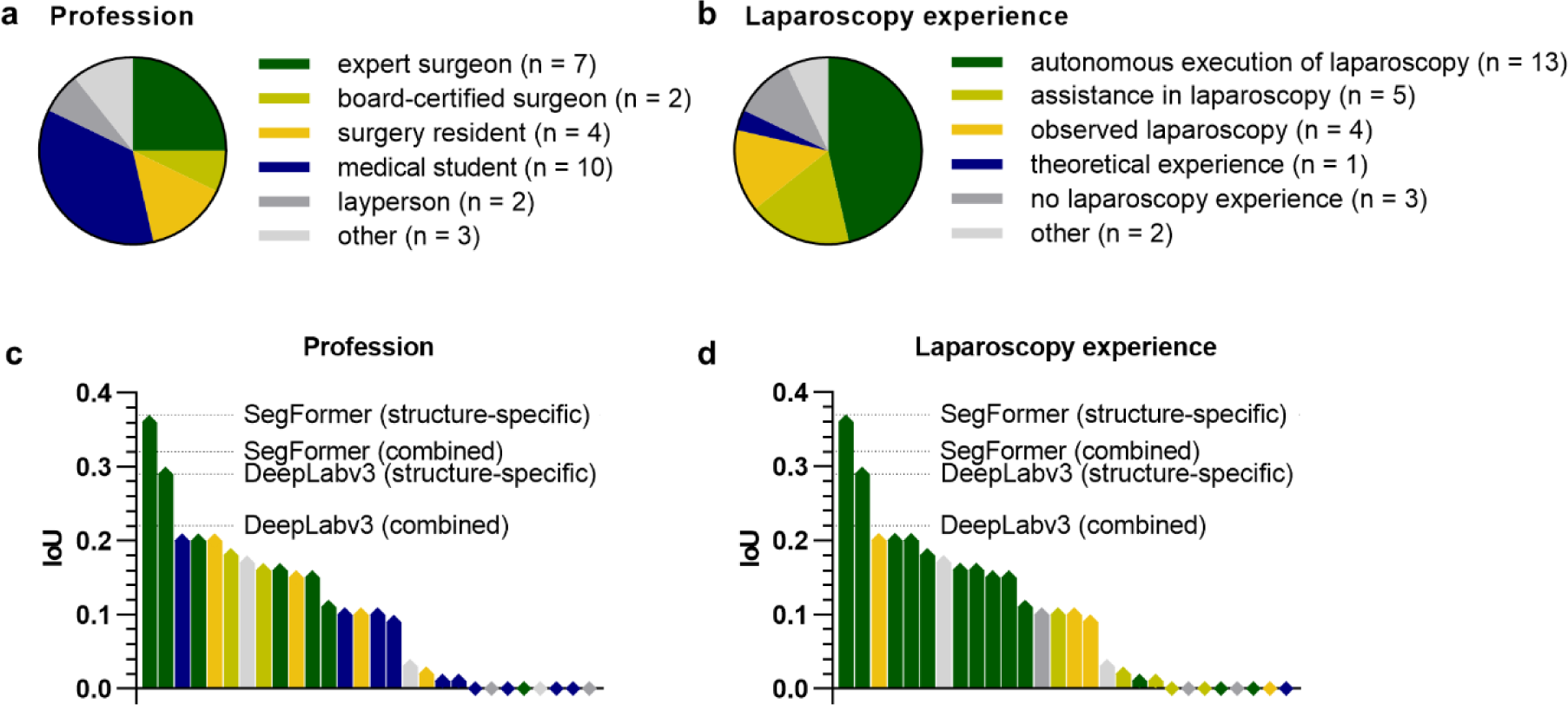
Comparison of pancreas segmentation performance of the structure-specific and the combined semantic segmentation models with a cohort of 28 human participants. **(a)** Distribution of medical and non-medical professions among human participants. **(b)** Distribution of laparoscopy experience among human participants. **(c)** Waterfall chart displaying the average pancreas segmentation IoUs of participants with different professions as compared to the IoU generated by the structure-specific and the combined semantic segmentation models. **(d)** Waterfall chart displaying the average pancreas segmentation IoUs of participants with varying laparoscopy experience as compared to the IoU generated by the structure-specific and the combined semantic segmentation models.

Comparing bounding box segmentations of the pancreas of human annotators, the medical and laparoscopy-specific experience of participants was mirrored by the respective IoUs describing the overlap between the pancreas annotation and the ground truth. The pancreas-specific segmentation models based on DeepLabv3 (IoU: 0.29) and SegFormer (IoU: 0.37) as well as the combined segmentation models based on DeepLabv3 (IoU: 0.21) and SegFormer (IoU: 0.32) outperformed at least 26 out of the 28 human participants (Figures 5 c and d). Overall, these results demonstrate that the developed models have clinical potential to improve the recognition of vulnerable anatomical structures in laparoscopy.

## Discussion

In surgery, misinterpretation of visual cues can result in objectifiable errors with serious consequences (22). Machine learning models could augment identification of anatomical structures during minimally-invasive surgery and thereby contribute to a reduction of surgical risks. However, data scarcity and suboptimal dataset quality, among other factors, drastically restrict the clinical impact of applications in the field of surgical data science (17,33–37). Based on a robust public dataset providing 13195 laparoscopic images with segmentations of eleven intra-abdominal anatomical structures, this study explores the potential of machine learning for automated segmentation of these organs, and compares algorithmic segmentation quality to that of humans with varying experience in minimally-invasive abdominal surgery.

In summary, the presented findings suggest that machine learning-based segmentation of intraabdominal organs and anatomical structures is possible and has the potential to provide clinically valuable information. At an average runtime of 71 ms per image, corresponding to a frame rate of 14 frames per second, the combined DeepLabv3-based model would facilitate near-real-time identification of eleven anatomical structures. In contrast, the SegFormer-based model is further from real-time performance at a runtime of 102 ms per image, resulting in a frame rate of less than 10 frames per second. These runtimes mirror the performances of non-optimized versions of the models, which can be significantly improved using methods such as TensorRT from Nvidia. However, with respect to generalizability and robustness, we observed substantially more accurate segmentation performance of the SegFormer-based models as compared to the DeepLabv3-based models when deployed to an external conventional laparoscopic dataset. Moreover, the structure-specific models exhibited a lack in accuracy and anatomical coherence, which can be explained by their organ-specific training process.

Measured by classical metrics of overlap between segmentation and ground truth, predictions were, overall, better for large and similar-appearing organs such as the abdominal wall, the liver, the stomach, and the spleen as compared to smaller and more diverse-appearing organs such as the pancreas, the ureter, or vesicular glands. Furthermore, poor image quality (i.e., images blurred by camera movements, presence of blood or smoke in images) was linked to lower accuracy of machine learning-based segmentations. Consequently, it is likely that a better nominal performance of the machine learning models could be achieved through selection of images from early phases of the surgery, in which such perturbations are not present. However, we purposely did not exclude images with suboptimal image quality, as selection on image level would introduce bias and thereby limit applicability. In this context, selection on patient level and on image level is a common challenge in computer vision (38) that can lead to skewed reporting of outcomes and poor performance on real-world data (34). Overall, our findings on the influence of image quality on segmentation performance imply that computer vision studies in laparoscopy should be carefully interpreted taking representativity and potential selection of underlying training and validation data into consideration.

Measured by classical metrics of overlap (e.g., IoU, F1 score, precision, recall, specificity) that are commonly used to evaluate segmentation performance, the structure-specific models and the combined models provided comparable segmentation performances on the internal test dataset. Interpretation of such metrics of overlap, however, represents a major challenge in computer vision applications in medical domains such as dermatology and endoscopy (39–41) as well as non-medical domains such as autonomous driving (42). In the specific use case of laparoscopic surgery, evidence suggests that such technical metrics alone are not sufficient to characterize the clinical potential and utility of segmentation algorithms (37,43). In this context, the subjective clinical utility of a bounding box-based detection system recognizing the common bile duct and the cystic duct at average precisions of 0.32 and 0.07, respectively, demonstrated by Tokuyasu *et al.*, supports this hypothesis (12). In colorectal surgery, anatomical misinterpretation during splenic flexure mobilization can result in iatrogenic lesions to the pancreas (24). In the presented analysis, the trained structure-specific and combined machine learning algorithms outperformed all human participants in the specific task of bounding box segmentation of the pancreas except for two surgical specialists with over 10 years of experience. This suggests that even for structures such as the pancreas with seemingly poor segmentation quality (segmentation IoU of the best-performing model: 0.37 ± 0.28 in the test set) have the potential to provide clinically valuable help in anatomy recognition. In this context, analysis of additional anatomical risk structures (i.e. ureters and blood vessels) and inclusion of more advanced personnel in future comparison studies will help better define the models’ capabilities in comparison with (expert) surgeons. Notably, the best average IoUs for pancreas segmentation achieved in this comparative study were 0.37 (for the SegFormer-based structure-specific model) and 0.36 (for the best human participant), which would both be considered less reliable segmentation quality measures on paper. This encourages further discussion about metrics for segmentation quality assessment in clinical AI. In the future, the potential of the described dataset (25) and organ segmentation algorithms could be exploited for educational purposes (44,45), for guidance systems facilitating real-time detection of risk and target structures (19,43,46,47), or as an auxiliary function integrated in more complex surgical assistance systems, such as guidance systems relying on automated liver registration (48).

The limitations of this work are mostly related to the dataset and general limitations of machine learning-based segmentation: First, the Dresden Surgical Anatomy Dataset is a monocentric dataset based on 32 robot-assisted rectal surgeries. Therefore, the images used for algorithm training and validation originate from one set of hardware and display organs from specific angles. As a consequence, given the lack of a laparoscopic image dataset with similarly rigorous organ annotations, generalizability and transferability of the presented findings to other centers and other minimally-invasive abdominal surgeries, particularly non-robotic procedures, could only be qualitatively investigated. Second, annotations were required for training of machine learning algorithms, potentially inducing some bias towards the way that organs were annotated in the resulting models. With respect to annotation quality, three individual annotations of each anatomical structure were reviewed by a single surgical expert. This represents a major limitation of the underlying dataset, which is reasoned in the time-consuming and effortful annotation process making the inclusion of more expert surgeons unfeasible. Given that annotations were based on specific annotation protocols including images (49) and all annotators had a medical background with several years of experience in the field of human anatomy (25), the quality of annotations can be considered high, despite the limited experience of the reviewing surgeon (4 years of experience in robot-assisted rectal surgery). This is particularly true when comparing the underlying dataset with other datasets commonly used in surgical data science that are often based on single annotations carried out by individuals without domain knowledge (17,26,50). Still, the way that organs are annotated may differ from individual healthcare professionals’ way of recognizing an organ. This is particularly relevant for organs such as the ureters or the pancreas, which often appear covered by layers of tissue. Here, computer vision-based algorithms that solely consider the laparoscopic images provided by the Dresden Surgical Anatomy Dataset for identification of risk structures will only be able to identify an organ once it is visible. For an earlier recognition of such hidden risk structures, more training data with meaningful annotations would be necessary. Importantly, the presented comparison to human performance focused on segmentation of visible anatomy as well, neglecting that humans (and possibly computers, too) could already identify a risk structure hidden underneath layers of tissue. Third, the dataset only includes individual annotated images. In some structures such as the ureter, video data offers considerably more information than still image data. In this context, it is conceivable that an incorporation of temporal aspects could result in major improvements of both human and algorithm recognition performance.

While the presented machine learning models show promise in improving the identification of anatomical structures in laparoscopy, their clinical utility still needs to be explored. Successful adoption of new technologies in surgery depends on factors beyond segmentation performance, runtime and generalizability, such as visualization of intraoperative decision support (51), human-machine interaction (52), and interface design. Therefore, interdisciplinary collaboration is critical to better understand respective surgeon needs. Moreover, prospective trials are needed to determine the impact of these factors on clinical outcomes. The existing limitations notwithstanding, the presented study represents an important addition to the growing body of research on medical image analysis in laparoscopic surgery, particularly by linking technical metrics to human performance.

In conclusion, this study demonstrates that machine learning methods have the potential to provide clinically relevant near-real-time assistance in anatomy recognition in minimally-invasive surgery. This study is the first to use the recently published Dresden Surgical Anatomy Dataset, providing baseline algorithms for organ segmentation and evaluating the clinical relevance of such algorithms by introducing more clinically meaningful comparators beyond classical computer vision metrics. Future research should investigate other segmentation methods, the potential to integrate high-level anatomical knowledge into segmentation models (38), the transferability of these results to other surgical procedures, and the clinical impact of real-time surgical assistance systems and didactic applications based on automated segmentation algorithms. Furthermore, seeing the DeepLabv3-based models outperform the SegFormer-based models in terms of run-time, but lacking in accuracy and generalizability, future research could focus on combining the two, in order to harness the best of both worlds.

## Data Availability

The Dresden Surgical Anatomy Dataset is publicly available via the following link: https://www.nature.com/articles/s41597-022-01719-2. All other data generated and analyzed during the current study are available from the corresponding authors on reasonable request. To gain access, data requestors will need to sign a data access agreement.

## Acknowledgments

Assistance with the study

The authors gratefully acknowledge excellent project coordination by Dr. Elisabeth Fischermeier and Dr. Grit Krause-Jüttler.

## Presentation

None.

## Abbreviations

AI: Artificial Intelligence
IoU: Intersection-over-Union
SD: Standard deviation

## Author contributions

FRK, JW, MD, SS, and SB conceptualized the study. FRK, FMR, and MC collected and annotated clinical and video data and contributed to data analysis. ACJ, SK, SL, and SB implemented and trained the neural networks and contributed to data analysis. JW, MD, and SS supervised the project, provided infrastructure and gave important scientific input. FRK drafted the initial manuscript text. All authors reviewed, edited, and approved the final manuscript.

