## Supplementary Material for "Anatomy Segmentation in Laparoscopic Surgery: Comparison of Machine Learning and Human Expertise – An Experimental Study"

**Table of Contents**

**Supplementary Figure 1**: Intersection-over-Union (IoU) ..………………………………………… **2**

**Supplementary Figure 2**: Pixel-wise organ segmentation with SegFormer-based structure-specific models trained on the respective organ subsets of the Dresden Surgical Anatomy Dataset ……………………………………………………………...………………………... **3**

**Supplementary Figure 3**: Pixel-wise organ segmentation with SegFormer-based combined model trained on the Dresden Surgical Anatomy Dataset across anatomical structure classes with a common encoder and structure-specific decoders ………………………………………….. **5**

**Supplementary Table 1**: Summary of performance metrics for anatomical structure segmentation using structure-specific models……………………………………………………….. **7**

**Supplementary Table 2**: Summary of performance metrics for anatomical structure segmentation using combined models (common encoder with structure-specific decoders)..…. **8**

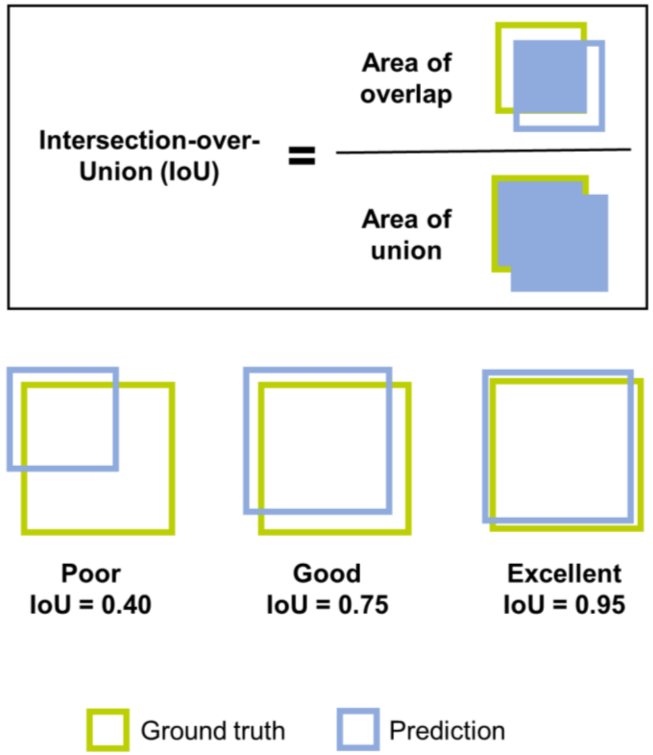


**Supplementary Figure 1: Intersection-over-Union (IoU).** Schematic illustration of calculation of IoU and examples of poor, good, and excellent IoU in segmentation tasks.

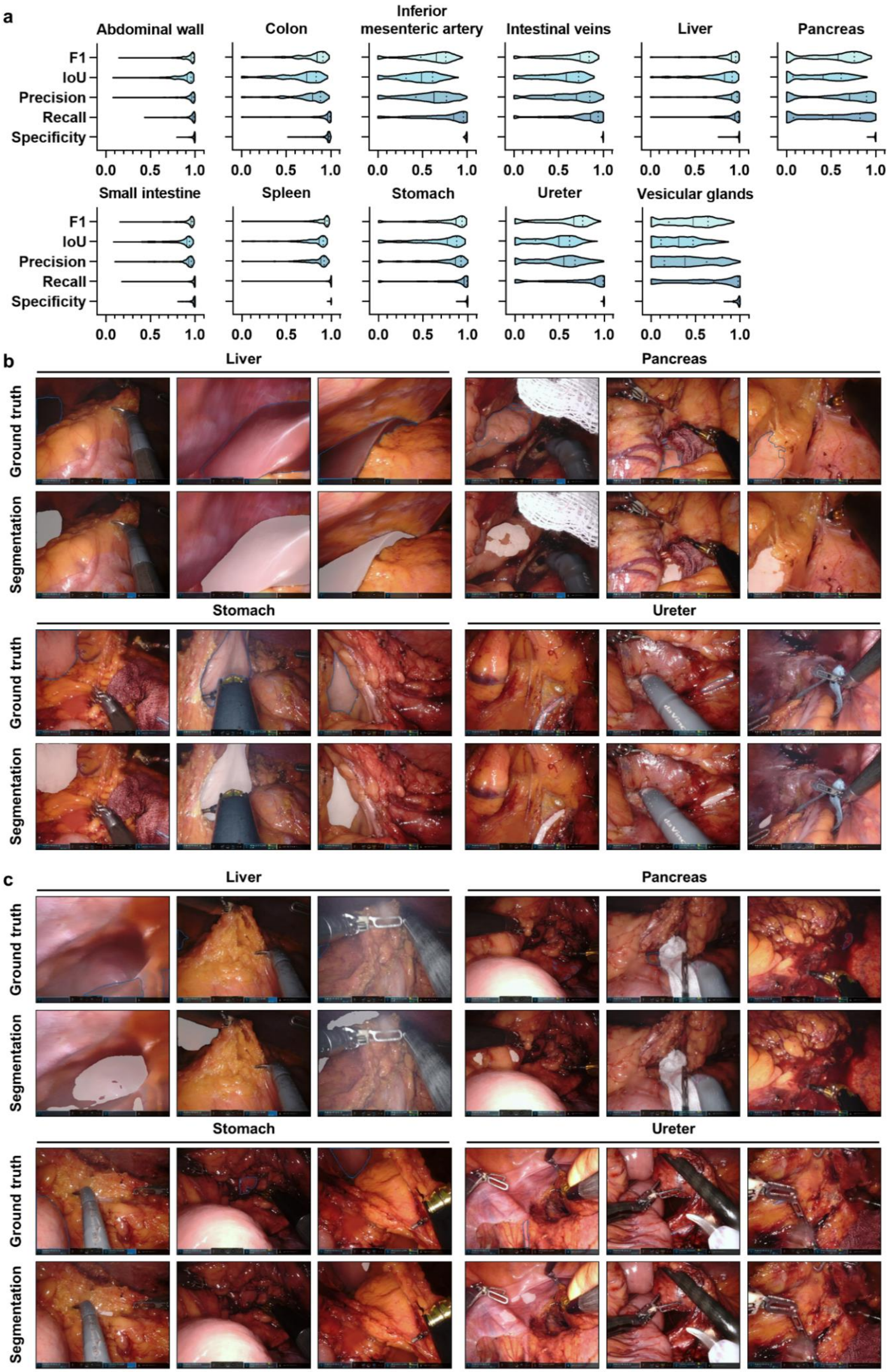
a
Abdominal wall
Colon
Inferior mesenteric artery
Intestinal veins
Liver
Pancreas
Small intestine
Spleen
Stomach
Ureter
Vesicular glands
F1
IoU
Precision
Recall
Specificity
0.0
0.5
1.0
b
Liver
Pancreas
Stomach
Ureter
Ground truth
Segmentation
c
Liver
Pancreas
Stomach
Ureter
Ground truth
Segmentation

**Supplementary Figure 2: Pixel-wise organ segmentation with SegFormer-based structure-specific models trained on the respective organ subsets of the Dresden Surgical Anatomy Dataset. (a)** Violin plot illustrations of performance metrics for SegFormer-based structure-specific segmentation models on the test dataset. The median and quartiles are illustrated as solid and dashed lines, respectively. **(b)** Example images from the test dataset with the highest IoUs for liver, pancreas, stomach, and ureter segmentation with SegFormer-based structure-specific segmentation models. Ground truth is displayed as blue line (upper panel), model segmentations are displayed as white overlay (lower panel). **(c)** Example images from the test dataset with the lowest IoUs for liver, pancreas, stomach, and ureter segmentation with SegFormer-based structure-specific segmentation models. Ground truth is displayed as blue line (upper panel), model segmentations are displayed as white overlay (lower panel).

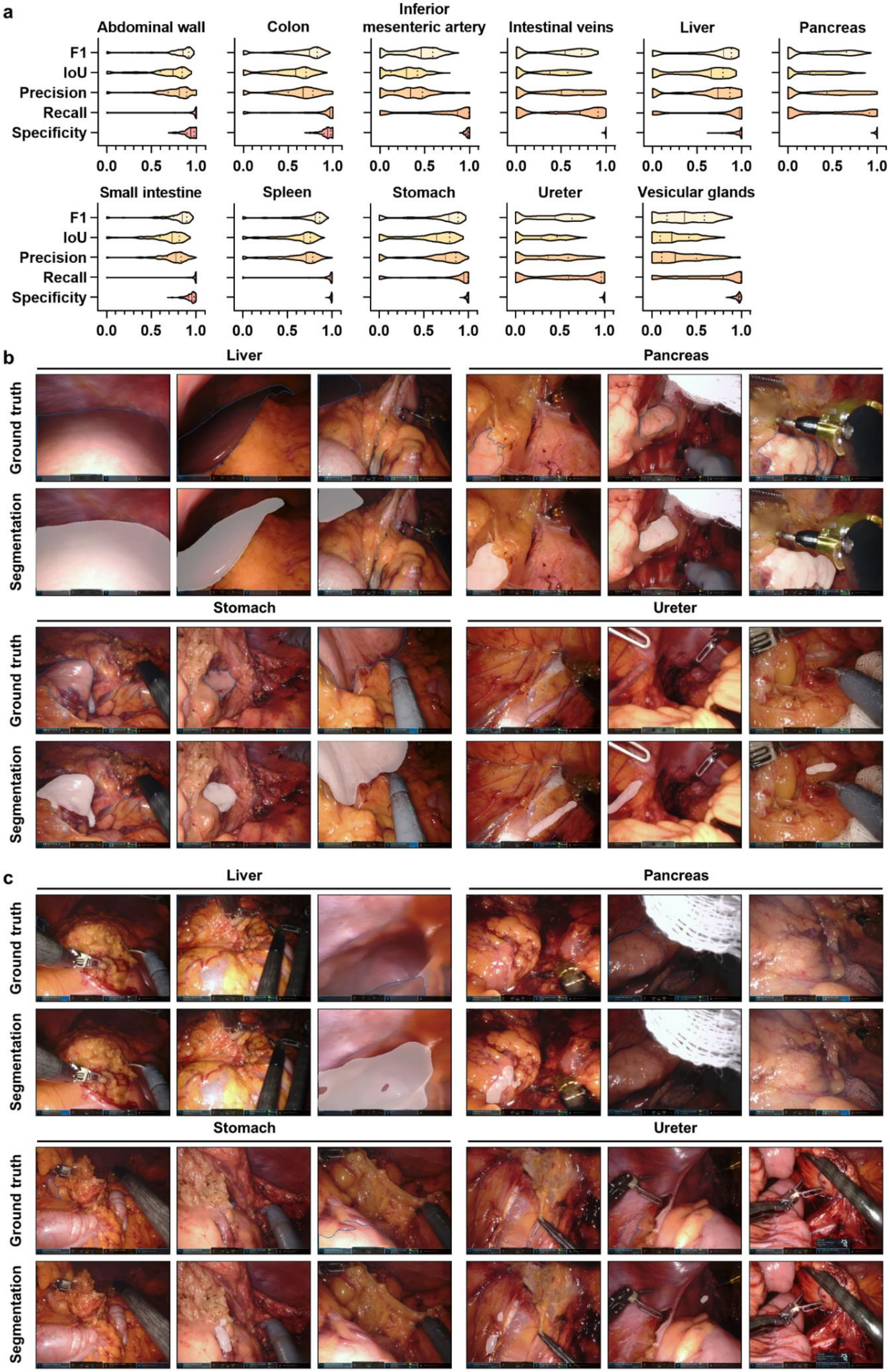
a
Abdominal wall
Colon
Inferior mesenteric artery
Intestinal veins
Liver
Pancreas
F1
IoU
Precision
Recall
Specificity
0.0 0.5 1.0
Small intestine
Spleen
Stomach
Ureter
Vesicular glands
b
Liver
Pancreas
Ground truth
Segmentation
Stomach
Ureter
c
Liver
Pancreas
Stomach
Ureter

**Supplementary Figure 3: Pixel-wise organ segmentation with SegFormer-based combined model trained on the Dresden Surgical Anatomy Dataset across anatomical structure classes with a common encoder and structure-specific decoders. (a)** Violin plot illustrations of performance metrics for the SegFormer-based combined segmentation model on the test dataset. The median and quartiles are illustrated as solid and dashed lines, respectively. **(b)** Example images from the test dataset with the highest IoUs for liver, pancreas, stomach, and ureter segmentation with the SegFormer-based combined segmentation model. Ground truth is displayed as blue line (upper panel), model segmentations are displayed as white overlay (lower panel). **(c)** Example images from the test dataset with the lowest IoUs for liver, pancreas, stomach, and ureter segmentation with the SegFormer-based combined segmentation model. Ground truth is displayed as blue line (upper panel), model segmentations are displayed as white overlay (lower panel).

**Supplementary Table 1: Summary of performance metrics for anatomical structure segmentation using structure-specific models.** Models were based on DeepLabv3 (a: training dataset, b: validation dataset) as well as SegFormer-based architectures (c: training dataset, d: validation dataset). For each metric, mean and standard deviation are displayed.

**a** DeepLabv3 (Training)

| Anatomical structure | F1 score | IoU | Precision | Recall | Specificity |
|---|---|---|---|---|---|
| Abdominal wall | 0.99 ± 0.01 | 0.97 ± 0.01 | 0.98 ± 0.01 | 0.99 ± 0.01 | 0.99 ± 0.00 |
| Colon | 0.97 ± 0.01 | 0.95 ± 0.03 | 0.95 ± 0.03 | 1.00 ± 0.00 | 0.99 ± 0.00 |
| Inferior mesenteric artery | 0.86 ± 0.06 | 0.76 ± 0.08 | 0.76 ± 0.08 | 1.00 ± 0.00 | 0.99 ± 0.00 |
| Intestinal veins | 0.87 ± 0.05 | 0.77 ± 0.08 | 0.77 ± 0.08 | 1.00 ± 0.00 | 1.00 ± 0.00 |
| Liver | 0.99 ± 0.02 | 0.97 ± 0.03 | 0.98 ± 0.02 | 1.00 ± 0.01 | 0.99 ± 0.01 |
| Pancreas | 0.80 ± 0.08 | 0.67 ± 0.10 | 0.67 ± 0.10 | 1.00 ± 0.00 | 0.99 ± 0.01 |
| Small intestine | 0.98 ± 0.01 | 0.96 ± 0.02 | 0.97 ± 0.02 | 1.00 ± 0.00 | 0.99 ± 0.00 |
| Spleen | 0.89 ± 0.08 | 0.82 ± 0.11 | 0.82 ± 0.11 | 1.00 ± 0.00 | 1.00 ± 0.00 |
| Stomach | 0.85 ± 0.10 | 0.76 ± 0.13 | 0.76 ± 0.13 | 1.00 ± 0.01 | 0.99 ± 0.01 |
| Ureter | 0.87 ± 0.07 | 0.77 ± 0.09 | 0.77 ± 0.09 | 1.00 ± 0.00 | 1.00 ± 0.00 |
| Vesicular glands | 0.68 ± 0.15 | 0.53 ± 0.16 | 0.53 ± 0.16 | 1.00 ± 0.04 | 0.99 ± 0.02 |

**b** DeepLabv3 (Validation)

| Anatomical structure | F1 score | IoU | Precision | Recall | Specificity |
|---|---|---|---|---|---|
| Abdominal wall | 0.91 ± 0.07 | 0.85 ± 0.11 | 0.92 ± 0.08 | 0.92 ± 0.10 | 0.97 ± 0.04 |
| Colon | 0.84 ± 0.14 | 0.74 ± 0.17 | 0.83 ± 0.16 | 0.87 ± 0.16 | 0.98 ± 0.02 |
| Inferior mesenteric artery | 0.62 ± 0.23 | 0.49 ± 0.22 | 0.63 ± 0.23 | 0.73 ± 0.27 | 0.99 ± 0.01 |
| Intestinal veins | 0.66 ± 0.25 | 0.53 ± 0.24 | 0.64 ± 0.25 | 0.75 ± 0.30 | 1.00 ± 0.00 |
| Liver | 0.75 ± 0.27 | 0.66 ± 0.28 | 0.83 ± 0.25 | 0.73 ± 0.29 | 0.98 ± 0.02 |
| Pancreas | 0.72 ± 0.16 | 0.58 ± 0.16 | 0.72 ± 0.14 | 0.77 ± 0.21 | 0.99 ± 0.01 |
| Small intestine | 0.88 ± 0.14 | 0.81 ± 0.18 | 0.88 ± 0.15 | 0.92 ± 0.14 | 0.97 ± 0.04 |
| Spleen | 0.74 ± 0.22 | 0.63 ± 0.23 | 0.72 ± 0.23 | 0.85 ± 0.21 | 0.99 ± 0.01 |
| Stomach | 0.64 ± 0.25 | 0.51 ± 0.24 | 0.58 ± 0.23 | 0.80 ± 0.28 | 0.97 ± 0.05 |
| Ureter | 0.45 ± 0.31 | 0.34 ± 0.25 | 0.57 ± 0.29 | 0.48 ± 0.36 | 1.00 ± 0.00 |
| Vesicular glands | 0.45 ± 0.25 | 0.32 ± 0.21 | 0.45 ± 0.28 | 0.65 ± 0.32 | 0.96 ± 0.04 |

**c** SegFormer (Training)

| Anatomical structure | F1 score | IoU | Precision | Recall | Specificity |
|---|---|---|---|---|---|
| Abdominal wall | 0.99 ± 0.01 | 0.98 ± 0.01 | 0.98 ± 0.01 | 1.00 ± 0.00 | 0.99 ± 0.00 |
| Colon | 0.89 ± 0.07 | 0.81 ± 0.10 | 0.82 ± 0.10 | 0.98 ± 0.02 | 0.98 ± 0.02 |
| Inferior mesenteric artery | 0.91 ± 0.04 | 0.83 ± 0.06 | 0.83 ± 0.06 | 1.00 ± 0.00 | 0.99 ± 0.00 |
| Intestinal veins | 0.84 ± 0.06 | 0.73 ± 0.09 | 0.73 ± 0.09 | 1.00 ± 0.00 | 1.00 ± 0.00 |
| Liver | 0.98 ± 0.02 | 0.97 ± 0.03 | 0.97 ± 0.03 | 1.00 ± 0.01 | 0.99 ± 0.01 |
| Pancreas | 0.83 ± 0.07 | 0.72 ± 0.09 | 0.72 ± 0.09 | 1.00 ± 0.00 | 0.99 ± 0.01 |
| Small intestine | 0.97 ± 0.01 | 0.94 ± 0.03 | 0.94 ± 0.03 | 1.00 ± 0.00 | 0.99 ± 0.01 |
| Spleen | 0.93 ± 0.05 | 0.87 ± 0.08 | 0.87 ± 0.08 | 1.00 ± 0.00 | 1.00 ± 0.00 |
| Stomach | 0.92 ± 0.06 | 0.86 ± 0.09 | 0.87 ± 0.09 | 1.00 ± 0.01 | 1.00 ± 0.00 |
| Ureter | 0.86 ± 0.07 | 0.77 ± 0.10 | 0.77 ± 0.10 | 1.00 ± 0.00 | 1.00 ± 0.00 |
| Vesicular glands | 0.67 ± 0.17 | 0.52 ± 0.18 | 0.53 ± 0.18 | 0.98 ± 0.08 | 0.98 ± 0.02 |

**d** SegFormer (Validation)

| Anatomical structure | F1 score | IoU | Precision | Recall | Specificity |
|---|---|---|---|---|---|
| Abdominal wall | 0.92 ± 0.09 | 0.85 ± 0.13 | 0.93 ± 0.08 | 0.92 ± 0.12 | 0.97 ± 0.04 |
| Colon | 0.81 ± 0.14 | 0.70 ± 0.17 | 0.75 ± 0.18 | 0.92 ± 0.13 | 0.96 ± 0.03 |
| Inferior mesenteric artery | 0.67 ± 0.22 | 0.54 ± 0.21 | 0.68 ± 0.24 | 0.76 ± 0.24 | 0.99 ± 0.01 |
| Intestinal veins | 0.70 ± 0.19 | 0.56 ± 0.20 | 0.64 ± 0.21 | 0.85 ± 0.21 | 1.00 ± 0.00 |
| Liver | 0.84 ± 0.21 | 0.76 ± 0.23 | 0.86 ± 0.22 | 0.85 ± 0.21 | 0.98 ± 0.03 |
| Pancreas | 0.76 ± 0.16 | 0.64 ± 0.17 | 0.77 ± 0.14 | 0.79 ± 0.20 | 0.99 ± 0.01 |
| Small intestine | 0.89 ± 0.13 | 0.83 ± 0.17 | 0.86 ± 0.16 | 0.96 ± 0.09 | 0.97 ± 0.04 |
| Spleen | 0.83 ± 0.17 | 0.73 ± 0.20 | 0.80 ± 0.19 | 0.91 ± 0.15 | 1.00 ± 0.01 |
| Stomach | 0.74 ± 0.25 | 0.64 ± 0.26 | 0.71 ± 0.26 | 0.84 ± 0.24 | 0.98 ± 0.03 |
| Ureter | 0.45 ± 0.33 | 0.35 ± 0.27 | 0.48 ± 0.32 | 0.54 ± 0.39 | 0.99 ± 0.01 |
| Vesicular glands | 0.49 ± 0.26 | 0.37 ± 0.23 | 0.49 ± 0.30 | 0.71 ± 0.32 | 0.96 ± 0.05 |

**Supplementary Table 2: Summary of performance metrics for anatomical structure segmentation using combined models (common encoder with structure-specific decoders).** Models were based on DeepLabv3 (a: training dataset, b: validation dataset) as well as SegFormer-based architectures (c: training dataset, d: validation dataset). For each metric, mean and standard deviation are displayed.

**a** DeepLabv3 (Training)

| Anatomical structure | F1 score | IoU | Precision | Recall | Specificity |
|---|---|---|---|---|---|
| Abdominal wall | 0.94 ± 0.03 | 0.89 ± 0.05 | 0.89 ± 0.05 | 1.00 ± 0.00 | 0.96 ± 0.02 |
| Colon | 0.89 ± 0.05 | 0.81 ± 0.08 | 0.81 ± 0.08 | 1.00 ± 0.01 | 0.97 ± 0.02 |
| Inferior mesenteric artery | 0.81 ± 0.10 | 0.69 ± 0.13 | 0.69 ± 0.13 | 0.99 ± 0.01 | 0.99 ± 0.01 |
| Intestinal veins | 0.75 ± 0.10 | 0.61 ± 0.12 | 0.61 ± 0.13 | 0.99 ± 0.03 | 0.99 ± 0.00 |
| Liver | 0.93 ± 0.06 | 0.88 ± 0.10 | 0.88 ± 0.10 | 1.00 ± 0.00 | 0.97 ± 0.03 |
| Pancreas | 0.75 ± 0.12 | 0.62 ± 0.14 | 0.63 ± 0.14 | 0.98 ± 0.03 | 0.99 ± 0.01 |
| Small intestine | 0.93 ± 0.04 | 0.87 ± 0.06 | 0.87 ± 0.06 | 1.00 ± 0.00 | 0.98 ± 0.02 |
| Spleen | 0.89 ± 0.09 | 0.81 ± 0.13 | 0.82 ± 0.13 | 0.98 ± 0.02 | 1.00 ± 0.00 |
| Stomach | 0.85 ± 0.10 | 0.75 ± 0.13 | 0.76 ± 0.13 | 1.00 ± 0.04 | 0.99 ± 0.01 |
| Ureter | 0.69 ± 0.14 | 0.54 ± 0.15 | 0.55 ± 0.15 | 0.99 ± 0.04 | 0.99 ± 0.01 |
| Vesicular glands | 0.77 ± 0.15 | 0.65 ± 0.17 | 0.67 ± 0.18 | 0.96 ± 0.11 | 0.99 ± 0.00 |

**b** DeepLabv3 (Validation)

| Anatomical structure | F1 score | IoU | Precision | Recall | Specificity |
|---|---|---|---|---|---|
| Abdominal wall | 0.87 ± 0.10 | 0.78 ± 0.14 | 0.85 ± 0.12 | 0.92 ± 0.13 | 0.94 ± 0.05 |
| Colon | 0.75 ± 0.17 | 0.62 ± 0.19 | 0.71 ± 0.18 | 0.87 ± 0.20 | 0.96 ± 0.03 |
| Inferior mesenteric artery | 0.52 ± 0.29 | 0.40 ± 0.25 | 0.57 ± 0.26 | 0.61 ± 0.35 | 0.99 ± 0.01 |
| Intestinal veins | 0.61 ± 0.25 | 0.48 ± 0.23 | 0.61 ± 0.22 | 0.72 ± 0.31 | 1.00 ± 0.00 |
| Liver | 0.59 ± 0.33 | 0.49 ± 0.32 | 0.70 ± 0.30 | 0.59 ± 0.37 | 0.97 ± 0.03 |
| Pancreas | 0.63 ± 0.21 | 0.49 ± 0.19 | 0.65 ± 0.23 | 0.70 ± 0.23 | 0.99 ± 0.01 |
| Small intestine | 0.84 ± 0.15 | 0.74 ± 0.18 | 0.80 ± 0.16 | 0.91 ± 0.17 | 0.96 ± 0.04 |
| Spleen | 0.68 ± 0.28 | 0.57 ± 0.28 | 0.69 ± 0.24 | 0.78 ± 0.32 | 0.99 ± 0.01 |
| Stomach | 0.46 ± 0.35 | 0.37 ± 0.31 | 0.62 ± 0.30 | 0.48 ± 0.39 | 0.99 ± 0.02 |
| Ureter | 0.30 ± 0.31 | 0.22 ± 0.24 | 0.52 ± 0.30 | 0.36 ± 0.39 | 1.00 ± 0.01 |
| Vesicular glands | 0.45 ± 0.24 | 0.32 ± 0.20 | 0.53 ± 0.29 | 0.51 ± 0.31 | 0.98 ± 0.02 |

**c** SegFormer (Training)

| Anatomical structure | F1 score | IoU | Precision | Recall | Specificity |
|---|---|---|---|---|---|
| Abdominal wall | 0.87 ± 0.10 | 0.78 ± 0.12 | 0.80 ± 0.11 | 0.97 ± 0.11 | 0.90 ± 0.08 |
| Colon | 0.78 ± 0.12 | 0.65 ± 0.14 | 0.65 ± 0.14 | 0.99 ± 0.08 | 0.94 ± 0.05 |
| Inferior mesenteric artery | 0.57 ± 0.15 | 0.41 ± 0.14 | 0.42 ± 0.15 | 0.97 ± 0.08 | 0.96 ± 0.02 |
| Intestinal veins | 0.61 ± 0.16 | 0.46 ± 0.16 | 0.48 ± 0.17 | 0.94 ± 0.14 | 0.99 ± 0.01 |
| Liver | 0.87 ± 0.10 | 0.78 ± 0.12 | 0.79 ± 0.11 | 0.99 ± 0.07 | 0.91 ± 0.08 |
| Pancreas | 0.56 ± 0.15 | 0.40 ± 0.14 | 0.42 ± 0.15 | 0.95 ± 0.13 | 0.98 ± 0.02 |
| Small intestine | 0.83 ± 0.08 | 0.72 ± 0.11 | 0.72 ± 0.11 | 0.99 ± 0.04 | 0.93 ± 0.06 |
| Spleen | 0.81 ± 0.16 | 0.70 ± 0.19 | 0.74 ± 0.17 | 0.93 ± 0.14 | 0.99 ± 0.01 |
| Stomach | 0.80 ± 0.14 | 0.69 ± 0.17 | 0.71 ± 0.16 | 0.97 ± 0.10 | 0.99 ± 0.01 |
| Ureter | 0.58 ± 0.22 | 0.44 ± 0.20 | 0.48 ± 0.20 | 0.90 ± 0.22 | 0.99 ± 0.01 |
| Vesicular glands | 0.54 ± 0.22 | 0.40 ± 0.21 | 0.41 ± 0.22 | 0.96 ± 0.11 | 0.98 ± 0.02 |

**d** SegFormer (Validation)

| Anatomical structure | F1 score | IoU | Precision | Recall | Specificity |
|---|---|---|---|---|---|
| Abdominal wall | 0.80 ± 0.17 | 0.69 ± 0.19 | 0.76 ± 0.14 | 0.90 ± 0.21 | 0.89 ± 0.07 |
| Colon | 0.69 ± 0.21 | 0.57 ± 0.22 | 0.65 ± 0.20 | 0.86 ± 0.25 | 0.94 ± 0.05 |
| Inferior mesenteric artery | 0.44 ± 0.27 | 0.32 ± 0.22 | 0.43 ± 0.24 | 0.68 ± 0.38 | 0.97 ± 0.02 |
| Intestinal veins | 0.57 ± 0.23 | 0.43 ± 0.21 | 0.53 ± 0.20 | 0.77 ± 0.30 | 0.99 ± 0.01 |
| Liver | 0.52 ± 0.36 | 0.43 ± 0.33 | 0.71 ± 0.28 | 0.57 ± 0.43 | 0.96 ± 0.06 |
| Pancreas | 0.62 ± 0.18 | 0.47 ± 0.17 | 0.56 ± 0.16 | 0.79 ± 0.24 | 0.98 ± 0.02 |
| Small intestine | 0.78 ± 0.21 | 0.67 ± 0.22 | 0.73 ± 0.17 | 0.91 ± 0.24 | 0.93 ± 0.06 |
| Spleen | 0.65 ± 0.30 | 0.55 ± 0.29 | 0.69 ± 0.26 | 0.75 ± 0.33 | 0.99 ± 0.01 |
| Stomach | 0.49 ± 0.37 | 0.40 ± 0.33 | 0.65 ± 0.29 | 0.57 ± 0.42 | 0.98 ± 0.02 |
| Ureter | 0.32 ± 0.30 | 0.24 ± 0.24 | 0.45 ± 0.29 | 0.44 ± 0.41 | 0.99 ± 0.01 |
| Vesicular glands | 0.51 ± 0.26 | 0.38 ± 0.23 | 0.47 ± 0.28 | 0.74 ± 0.29 | 0.96 ± 0.04 |
